# Annotation and Information Extraction of a Consumer-Friendly Health Website for Enhancing Laboratory Test Reporting in Patient Portals

**DOI:** 10.1101/2022.12.19.22283692

**Authors:** Zhe He, Shubo Tian, Arslan Erdengasileng, Karim Hanna, Yang Gong, Zhan Zhang, Xiao Luo, Mia Liza A. Lustria

## Abstract

Viewing laboratory test results is patients’ most frequent activity when accessing patient portals, but lab results can be very confusing for patients. Previous research has explored various ways to present lab results, but few have attempted to provide tailored information support based on individual patient’s medical context. In this study, we collected and annotated interpretations of textual lab result in 251 health articles about laboratory tests from AHealthyMe.com. Then we evaluated transformer-based language models including BioBERT, ClinicalBERT, RoBERTa, and PubMedBERT for recognizing key terms and their types. Using BioPortal’s term search API, we mapped the annotated terms to concepts in major controlled terminologies. Results showed that PubMedBERT achieved the best F1 on both strict and lenient matching criteria. SNOMED CT had the best coverage of the terms, followed by LOINC and ICD-10-CM. This work lays the foundation for enhancing the presentation of lab results in patient portals by providing patients with contextualized interpretations of their lab results and individualized question prompts that they can, in turn, refer to during physician consults.

## Introduction

With the wide adoption of EHR systems, more patients have direct access to their clinical data via patient portals, allowing them to view their visit summaries, lab test results, medications, allergies, diagnoses, etc. [1]. Research shows that giving patients access to their medical records through patient portals improves health behaviors, medication adherence, and self-management of chronic conditions; enhances doctor-patient communication; reduces utilization of high-cost healthcare services among patients with chronic conditions; improves recovery; reduces hospital readmissions; and facilitates timely and patient-centered care [2,3]. Viewing lab test results is the most frequent activity patients do when accessing patient portals but lab results can be very confusing for patients [1]. Most patient portals present lab results in tabular format with a universal reference range, similar to the format seen by clinicians [5,6].

Merely providing access to their records is insufficient for improving patient engagement in their care because many patients, especially those with low health literacy, cannot make sense of the results and act upon them [4]. Patients with limited health literacy are more likely to misinterpret or misunderstand their lab results, which in turn, may delay their seeking critical medical attention [1,9]. Various studies have found a significant inverse relationship between health literacy and numeracy and the ability to make sense of lab results [7,8]. Giardina et al. [10] conducted interviews with 93 patients and found that nearly two-thirds did not receive any explanation for their lab results, and 46% conducted online searches to understand their results better. Similarly, another study found that patients who were unable to assess the gravity of their test results were more likely to wait for their doctor to call or seek information on the Internet [8].

As such, there is a pressing need to study how to present lab test results to improve patients’ understanding and to support shared decision making. Various studies, including our own, have explored different strategies for presenting numerical data to patients, to name a few: use of reference ranges, tables, charts, color, text, numerical data with verbal explanations, etc. [5,11,12]. Nonetheless, to the best of our knowledge, no studies have explored how to provide tailored textual explanations based on the medical context of individual patients. There are abundant online health sources that provide lab test information. However, they are not organized in a way that is computable. With informatics approaches such as natural language processing and ontologies, we can build a robust knowledge base that systematically organizes relevant content from online sources to enable flexible querying and linkage to patients’ medical records. Consequently, this system will be able to facilitate the generation of tailored support.

In this study, we demonstrate the feasibility of extracting contextualized interpretations of lab test results from credible health articles collected from AHealthyMe.com. We employed a multi-stage annotation procedure to annotate key terms, entity types, and their relationships in the textual content of collected records. To support large-scale information extraction, we evaluated transformer-based language models for recognizing key terms and their entity types. We also used BioPortal’s term search function to map the annotated terms to concepts from controlled vocabularies, including SNOMED CT, LOINC, RxNORM, and ICD10-CM to improve interoperability with other sources and systems. The contributions of this work are two-fold. First, we provide an annotation dataset of interpretations of lab results from over 200 records of health articles from a credible health source. Second, we demonstrate the feasibility of building a computable knowledge base using entity recognition, entity linking, and knowledge graph techniques. Such a knowledge base can be used to enhance the presentation of lab results in patient portals and, in turn, aid patients in understanding their lab results and help them prepare for follow-up consults with their doctors.

## Related Work

### Challenges for Lab Result Comprehension

Previous studies have investigated the challenges associated with lab results comprehension. Zhang et al. [11] used both a web-based survey and semi-structured interviews to understand patient challenges and needs in comprehending lab test results. They found that patients need both generic information and tailored information such as treatment options, prognosis, and action. They also found that result normality, health literacy, and technology proficiency significantly impact people’s perceptions of the use of patient portals to view and interpret laboratory results. In another study, Zhang et al. [13] analyzed questions related to lab tests posted on a social Q&A site and found that most patients need support for understanding their test results, doctor’s diagnoses, learning about lab tests, and figuring out the next steps. Zikmund-Fisher et al. [7] conducted an online survey to determine how numeracy level (an aspect of health literacy) affects individuals’ comprehension of lab results. Notably, only slightly half (51.24%) of the participants could correctly identify out-of-range hemoglobin readings. Compared to those with lower literacy, those with higher numeracy had greater sensitivity for out-of-range results and showed more initiative with contacting doctors. Providing support for patients with low health literacy and numeracy through the use of interpretation-based approaches can help improve their ability to correctly identify abnormal lab test values as well as their ability to participate more actively in managing their health.

### Improving Lab Results Comprehension

Existing work have also investigated ways to improve the comprehension of lab results. Kopanitsa [14] studied how patients perceive interpretations of lab results automatically generated by a clinical decision support system. They found that all the patients who received interpretations of the abnormal test results had a significantly higher rates of follow-up (71%) compared to those who received only test results without interpretations (49%). Patients appreciate the timeliness of automatically-generated interpretations compared to interpretations they can receive from a doctor. Zikmund-Fisher et al. [16] surveyed 1,618 adults in the US to assess how different visual presentations of lab results influence their perceived urgency. They found that a visual line display -- that included both the standard range and a harm anchor reference point which many doctors may not consider as particularly concerning -- reduced the perceived urgency of close-to-normal alanine aminotransferase and creatinine results (p-value < .001). In a similar study, Scherer et al. [17] tested different display formats of HbA1c result (i.e., a table, a simple line, and a number line with diagnostic categories indicated via colored blocks). They found that the goal range only displays achieved higher levels of comprehension of test results and decreased negative reaction compared with the no goal range and goal range added conditions. They concluded that replacing the standard range with a clinically appropriate goal range could improve the comprehension of lab results. Morrow et al. [18] investigated whether providing verbally-, graphically-, and video-enhanced context for patient portal messages about lab results can improve responses to the messages. They found that compared to a standardized format verbally- and video-enhanced contexts were able to improve older adults’ gist but not verbatim memory. All of these aforementioned studies focused on improving lab results reporting but the approaches used did not take individual patients’ health status into account.

## Methods

### Overall Workflow

**Figure 1** shows the overall workflow of this study. After we obtained all the health articles about laboratory tests from AHealthyMe.com, we extracted the sections about lab results interpretation. Following a rigorous multi-stage annotation and curation process, we generated an annotated dataset of over 200 records of lab result interpretations. Then we trained and tested transformer-based models for named entity recognition of the key terms and used BioPortal’s term search API to map the annotated terms to concepts in well-established controlled terminologies. Lastly, we stored the annotation results in Neo4j to enable graph-based queries. The detail of the workflow is described below.

**Figure 1.**
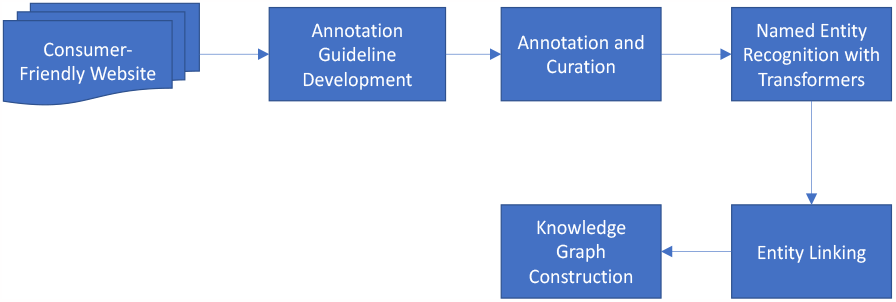
Overall workflow of this study.

### Data Source

AHealthyMe is a credible health website created and maintained by Blue Cross Blue Shield of Massachusetts. It provides a health library with articles, decision tools, and symptom checker on a wide range of health and wellness topics. The health articles are reviewed and verified by medical professionals. The website provides consumer-oriented semi-structured content about lab tests organized under eight questions: “*Does this test have other names?*”, “*What is this test?*”, “*Why do I need this test?*”, “*What other tests might I have along with this test?*”, “*What do my test results mean?*”, “*How is this test done?*”, “*Does this test pose any risks?*”, “*What might affect my test results?*”, and “*How do I get ready for this test?*”. We crawled the webpages of “*Tests & Procedures*” category of AHealthyMe.com and obtained 251 health articles about lab tests. Then we extracted the sections “*Does this test have other names?*” and “*What do my test results mean?*” from these articles to create the corpus of our study and combine these sections as a single record for a lab test. We focused on these two sections because they contain primary interpretations about lab results. **Figure 2** shows an example of a lab result interpretation for hemoglobin.

**Figure 2.**
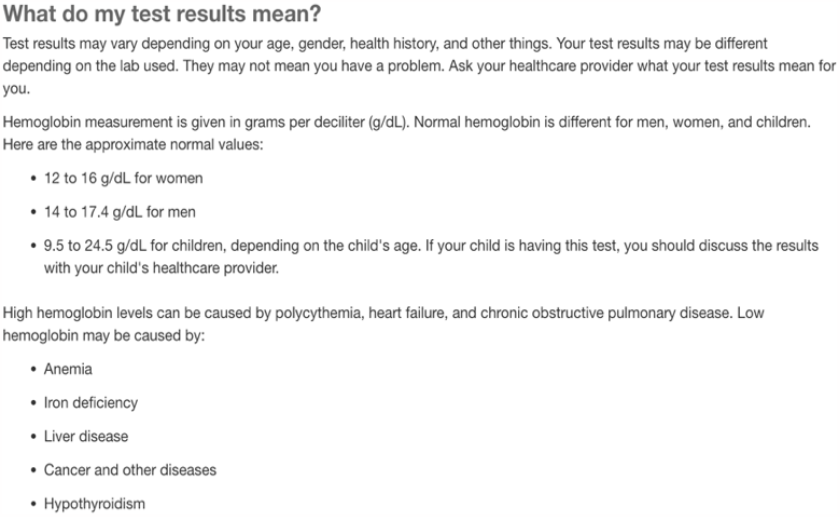
The section of lab result interpretation for hemoglobin.

### Annotation Process

To identify key information from the records, we annotated the data using a web-based annotation tool called INCEpTION, a semantic annotation platform that provides intelligent annotation assistance and knowledge management. It supports the seamless collaboration of multiple annotators, curators, and managers to ensure the annotation quality. We followed a rigorous process consisting of five steps: (1) annotation guideline development, (2) a pilot test with training, inter-rater reliability assessment, and conflict resolution, (3) separate annotation, (4) curation, (5) quality assurance. In Step 1, PI ZH and the doctoral student AE annotated 37 samples randomly selected from all the records and identified 12 entity types (e.g., “lab name”, “normal range”, “indication”, “condition”) and 7 relationships between these entities. PI ZH developed the annotation guideline that explains each entity type and relationship with examples. The annotation guideline was further reviewed and verified by AE. The explanation about the entity types with examples are provided in **Table 1**. Note that “normal_range” and “abnormal range” labels are used for both numeric value ranges and binary test results (e.g., positive, negative). The details about the relationships between the entities are provided in **Table 2. Figure 3** shows the relationships between the entities. In Step 2, four undergraduate pre-med students were recruited to annotate the same set of 37 samples following the guideline. PI had weekly meetings with the students to go over the annotated examples and made clarifications. Interrater reliability was calculated between each student annotator and the PI. Conflicts and inconsistencies were resolved after discussion. In Step 3, four students separately annotated all the 251 records. Each record was annotated by two annotators. In Step 4, PI and AE performed the curation and consolidated the annotation. In Step 5, we performed quality assurance of the annotations with aggregate analysis of both entity types and relationships and made corrections to the annotations. **Figure 4** shows the number of relationships between the entities in the annotated dataset.

**Table 1.** Entity types and example annotations.

| Entity Type (#) | Explanation | Example Annotation (bold texts were annotated) |
| --- | --- | --- |
| indication (N=730) | indication of a normal or abnormal value range or result | A1C from 5.7% to 6.4%. **You may have prediabetes.** This means **you have a higher risk for diabetes in the future.** A1C of 6.5% or above on 2 separate tests. **You may have diabetes.** |
| alt_lab_name (N=620) | alternative name of a lab test | Creatinine (**Serum creatinine; blood creatinine**): 0.9 to 1.3 mg/dL for adult males, 0.6 to 1.1 mg/dL for adult females, 0.5 to 1.0 mg/dL for children ages 3 to 18 years. 0.3 to 0.7 mg/dL for children younger than age 3. |
| abnormal_range (N=406) | a value range or binary test result that is abnormal. For value range, it may include the value(s), preposition(s), comparison operator(s). For test result, it may be positive or negative. | A1C from **5.7% to 6.4%**. You may have prediabetes. This means you have a higher risk for diabetes in the future. A1C of **6.5% or above** on 2 separate tests. You may have diabetes. |
| lab_name (N= 353) | the name of the lab test (e.g., “Creatinine”, “Hemoglobin”) | **Creatinine** (Serum creatinine; blood creatinine): 0.9 to 1.3 mg/dL for adult males, 0.6 to 1.1 mg/dL for adult females, 0.5 to 1.0 mg/dL for children ages 3 to 18 years. 0.3 to 0.7 mg/dL for children younger than age 3. |
| normal_range (N=350) | normal value range or binary result of a lab test | Creatinine (Serum creatinine; blood creatinine): **0.9 to 1.3 mg/dL** for adult males, **0.6 to 1.1 mg/dL** for adult females, **0.5 to 1.0 mg/dL** for children ages 3 to 18 years. **0.3 to 0.7 mg/dL** for children younger than age 3. |
| specimen_type (N=118) | the specimen type of a lab test (e.g., urine, blood) | Albumin (**Urine**) (Urine albumin, 24-hour urine test for albumin) |
| condition (N=95) | a certain condition on which a certain result is dependent | Creatinine (Serum creatinine; blood creatinine): 0.9 to 1.3 mg/dL for **adult males**, 0.6 to 1.1 mg/dL for **adult females**. |
| cause (N=91) | cause of a certain abnormal range/result | A lower-than-normal level of protein C may be caused by: **Blood-thinning medicines, such as warfarin, Kidney problems, Deficiency in vitamin K, inherited protein C deficiency, Condition that causes the blood to clot too much (consumptive coagulopathy)** |
| age_group (N=76) | age group can be an age range (e.g., “younger than 50”), a certain population group defined by age (e.g., adults, newborn, children) | Creatinine (Serum creatinine; blood creatinine): 0.9 to 1.3 mg/dL for adult males, 0.6 to 1.1 mg/dL for adult females, 0.5 to 1.0 mg/dL for **children ages 3 to 18 years**. 0.3 to 0.7 mg/dL for **children younger than age 3**. |
| action (N=71) | action to take to achieve a goal or given a certain result | To lower the calcium level in your urine, your healthcare provider might suggest that **you eat more vegetables and fruits and less animal products, like red meat and eggs.** |
| gender (N=54) | male or female | Creatinine (Serum creatinine; blood creatinine): 0.9 to 1.3 mg/dL for adult **males**, 0.6 to 1.1 mg/dL for adult **females**. |
| goal (N=2) | a goal to achieve | **To lower the calcium level in your urine**, your healthcare provider might suggest that you eat more vegetables and fruits and less animal products, like red meat and eggs. |

**Table 2.** Relationship types and example annotations.

| Relationship Type | Usage | Example |
| --- | --- | --- |
| has_indication (N=728) | the relation between “abnormal_range”/“normal_range” and “indication”. | “Hemoglobin A1c”<br>“from 5.7% to 6.4%” **has_indication** “You may have prediabetes. This means you have a higher risk for diabetes in the future” |
| has_range (N=714) | The relation between “lab_name” and “normal_range”/“abnormal_range” | “Hemoglobin A1c” **has_range** “below 5.7%” |
| has_alt_name (N=616) | the relation between “lab_name” and “alt_lab_name”. | “Hemoglobin A1c” **has_alt_name** “HbA1c” |
| has_condition (N=225) | the relation between “normal_range”/“abnormal range” and “condition” /“age_group”/“gender”. | “Creatinine”:<br>“0.9 to 1.3 mg/dL” **has_condition** “adult males” |
| has_specimen_type (N=112) | The relation between “lab_name” and “specimen_type”. | “Albumin” **has_speciment_type** “Urine” |
| caused_by (N=94) | the relation between “abnormal_range” and “cause”. | “lower-than-normal level of protein C” **caused_by** “Blood-thinning medicines, such as warfarin” |
| has_action (N=67) | the relation between “normal result/abnormal result/indication” or “goal” and “action”. | “Calcium”<br>“To lower the calcium level in your urine”<br>**has_action** “eat more vegetables and fruits and less animal products, like red meat and eggs.” |

**Figure 3.**
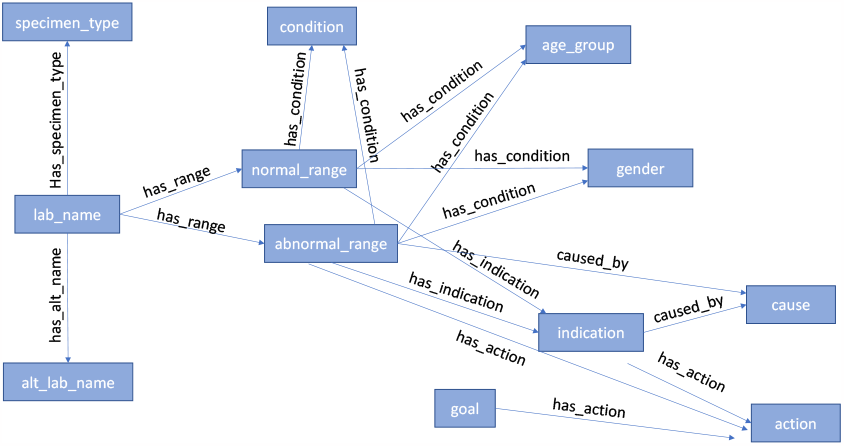
Relationships between entities

**Figure 4.**
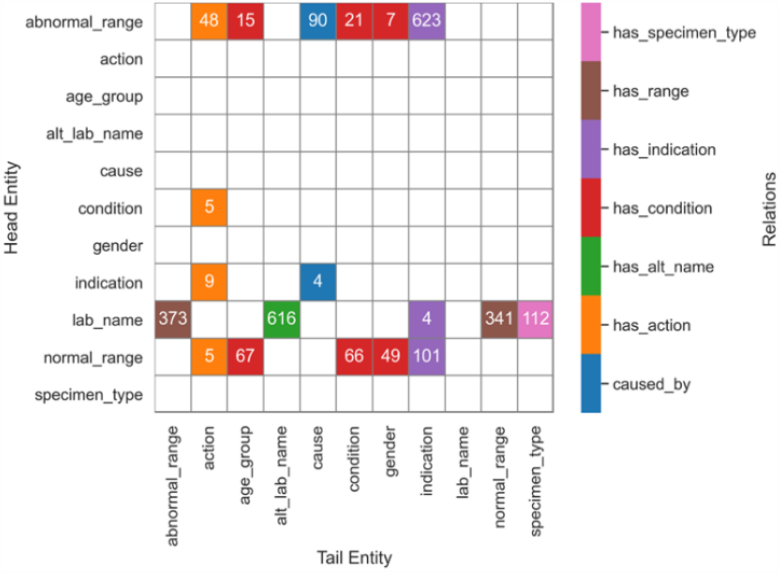
Number of annotated instances of relationships between entities.

### Named Entity Recognition (NER) with Transformers

Transformer models have achieved superior performance in many natural language processing tasks such as named entity recognition and sentiment analysis. These models pretrained with large corpus can be finetuned with annotated datasets for NER. To explore the feasibility of automatic extraction of key terms from lab result interpretations, we used our annotated dataset to finetune 4 widely used SOTA transformer-based models including BioBERT, ClinicalBERT, RoBERTa, and PubMedBERT for named entity recognition. These models are based on BERT model, a multilayer bidirectional transformer-based encoder model pretrained with BooksCorpus and English Wikipedia using masked language modeling and optimized by next sentence prediction [19]. BioBERT was generated by further pre-training BERT with PubMed abstracts (4.5 billion words) and PMC full-text articles (13.5 billion words) [20]. ClinicalBERT was generated by pretraining BERT with clinical notes in MIMIC III (0.5 billion words) [21]. RoBERTa has the same architecture as BERT and was pretrained with longer web content using dynamic masked language modeling which randomly selects spans of text to mask at each training epoch and different training strategies [22]. We further pre-trained RoBERTa model with clinical trial eligibility criteria corpus of ClinicalTrials.gov [23]. PubMedBERT was trained from scratch using PubMed abstracts [24]. We experimented with training the models with the entire paragraphs of lab result explanations and individual sentences. We converted the annotated corpus into BIO format (e.g., “B_lab_name”, “I_lab_name”, and “O” labels for the beginning, inside, and outside of a lab name term, respectively) using SpaCy and split the data into 70% training, 10% validation, and 20% testing (see **Table 4**). As such, the NER task is to predict the label for individual tokens. We evaluated model performance using precision, recall, and F1 based on both the strict matching criterion (i.e., exact match of both the entity type and entity with the annotated entity) and lenient matching criterion (i.e., requires the predicted entity overlaps with the annotated entity).

### Entity Linking

Modern EHR systems use controlled terminologies such as SNOMED CT, LOINC, ICD-9/ICD-10, RxNORM to encode medical information for patients. To support the construction of the knowledge base that allows linkage to EHR data, it is important to use concepts in well-established controlled vocabularies to encode the annotated entities. For entity linking, we employed the term search function of BioPortal, which allows us to identify terms from well-established controlled vocabularies that match the annotated entities from the dataset. The reason we could reliably use BioPortal is because we were able to limit the search of matching terms from controlled vocabularies that are relevant to a particular entity type. **Table 3** lists the entity type and the controlled vocabularies that were considered for term search.

**Table 3.** Entity types and relevant controlled vocabularies

| Entity Type | Controlled Vocabularies |
| --- | --- |
| lab_name | LOINC, SNOMED CT |
| alt_lab_name | LOINC, SNOMED CT |
| indication | ICD-10, SNOMED CT |
| specimen_type | SNOMED CT |
| cause | RxNORM, ICD-10, SNOMED CT |
| condition | SNOMED CT |
| age_group | SNOMED CT, LOINC |
| action | SNOMED CT, LOINC |
| gender | SNOMED CT, LOINC |

## Results

### Basic Characteristics of the Annotated Dataset

Of all the 251 lab test comprehension records, our systematic annotation discovered 2,964 annotated entities of 11 entity types and 2,556 relationships of 7 relation types. Among the 11 entity types, *Indication* entities appear the most frequently (N=730) followed by alt_lab_name entities (N=620). Entities of gender appear the least frequently in the records, with a handful of 54 annotated entities. **Table 4** shows the number of instances for each entity type.

**Table 4.** Basic characteristics of the paragraphs, sentences and annotated terms.

| Dataset and Data Split | Train | Validation | Test | Total |
| --- | --- | --- | --- | --- |
| Paragraphs | 175 | 25 | 51 | 251 |
| Sentences | 1,286 | 143 | 331 | 1,760 |
| Annotated terms | 2,066 | 277 | 621 | 2,964 |
| indication | 542 | 51 | 137 | 730 |
| alt_lab_name | 400 | 60 | 160 | 620 |
| abnormal_range | 301 | 33 | 72 | 406 |
| lab_name | 247 | 29 | 77 | 353 |
| normal_range | 250 | 34 | 66 | 350 |
| specimen_type | 75 | 10 | 33 | 118 |
| condition | 70 | 8 | 17 | 95 |
| cause | 46 | 27 | 18 | 91 |
| age_group | 47 | 13 | 16 | 76 |
| action | 51 | 4 | 16 | 71 |
| gender | 37 | 8 | 9 | 54 |

### Performance of Named Entity Recognition Models

As shown in **Table 5**, the best model for strict matching in terms of F1 is PubMedBERT (0.60), followed by RoBERTa_trial (0.59), ClinicalBERT (0.56), BioBERT (0.56). Best F1 score of lenient matching is PubMedBERT (0.82), followed by RoBERTa (0.81), ClinicalBERT (0.81), BioBERT (0.81). NER models with sentences as input had a slightly worse performance with the best models being BioBERT (0.57) for strict matching and RoBERTa for lenient matching (0.89). For brevity, the detailed results are not provided in Table 5.

**Table 5.** Performance of transformer-based NER models

| Model | Strict Matching |  |  | Lenient Matching |  |  |
| --- | --- | --- | --- | --- | --- | --- |
|  | Precision | Recall | F1 | Precision | Recall | F1 |
| **NER using paragraphs as input** |  |  |  |  |  |  |
| BioBERT | 0.5262 | 0.5974 | 0.5596 | 0.7844 | 0.8406 | 0.8115 |
| ClinicalBERT | 0.5281 | 0.6055 | 0.5641 | 0.7879 | 0.8374 | 0.8119 |
| RoBERTa | 0.5461 | 0.6393 | 0.5890 | 0.7799 | 0.8519 | 0.8143 |
| PubMedBERT | **0.5670** | **0.6409** | **0.6017** | **0.7949** | **0.8551** | **0.8239** |

As PubMedBERT has the overall best performance, we further analyzed its performance by entity types. As shown in **Table 6**, NER for entity types with more instances (i.e., alt_lab_name, speciment_type, age_group, gender, lab_name, normal_range, indication, abormal_range) achieved F1 over 0.7 using the lenient matching criterion. NER for entity types with fewer terms in our dataset (i.e., condition, cause, action) had F1 lower than 0.5.

**Table 6.** Performance of PubMedBERT NER models by entity types

| Entity Type | Strict Matching |  |  | Lenient Matching |  |  |
| --- | --- | --- | --- | --- | --- | --- |
|  | Precision | Recall | F1 | Precision | Recall | F1 |
| alt_lab_name | 0.9320 | 0.8562 | 0.8925 | 0.9932 | 0.9375 | 0.9645 |
| specimen_type | 0.8750 | 0.8485 | 0.8615 | 0.9062 | 0.8788 | 0.8923 |
| age_group | 0.7778 | 0.8750 | 0.8235 | 0.8889 | 0.9375 | 0.9125 |
| gender | 0.7273 | 0.8889 | 0.8000 | 0.8182 | 1.0000 | 0.9000 |
| lab_name | 0.6706 | 0.7403 | 0.7037 | 0.7765 | 0.8312 | 0.8029 |
| normal_range | 0.5897 | 0.6970 | 0.6389 | 0.8333 | 0.9394 | 0.8832 |
| indication | 0.3672 | 0.4745 | 0.4140 | 0.7458 | 0.8394 | 0.7898 |
| abnormal_range | 0.3500 | 0.4861 | 0.4070 | 0.6600 | 0.8889 | 0.7575 |
| condition | 0.3333 | 0.2353 | 0.2759 | 0.5000 | 0.3529 | 0.4138 |
| cause | 0.1429 | 0.1667 | 0.1538 | 0.6190 | 0.4444 | 0.5174 |
| action | 0.0476 | 0.0625 | 0.0541 | 0.4762 | 0.5625 | 0.5158 |
| **Overall** | **0.5670** | **0.6409** | **0.6017** | **0.7949** | **0.8551** | **0.8239** |

### Entity Linking Results

Table 7 shows the entity linking results by identifying the terms in major medical controlled terminologies that match the annotated entities in the dataset with BioPortal’s term search function. We present both the exact match and partial match results for each of the relevant terminologies and each entity type. Exact match means that the term in the controlled terminology is exactly the same as the annotated entity. Partial match means that the term in the controlled terminology is part of the annotated entity. Note that we only performed partial match for those terms that could not find a matching concept using exact match. SNOMED CT can cover 80.3% - 100% annotated entities when both exact match and partial match are counted.

**Table 7.** Coverage of the annotated entities by controlled vocabularies

| Entity Type | LOINC |  | SNOMED CT |  | RxNORM |  | ICD-10-CM |  | Total |  |
| --- | --- | --- | --- | --- | --- | --- | --- | --- | --- | --- |
|  | Exact | Partial | Exact | Partial | Exact | Partial | Exact | Partial | Exact | Partial |
| indication | -- |  | 21.6%<br>(158/730) | 65.6%<br>(479/730) | -- |  | 5.6%<br>(41/730) | 34.4%<br>(251/730) | 21.6%<br>(158/730) | 65.6%<br>(479/730) |
| alt_lab_name | 11.3%<br>(70/620) | 66.6%<br>(413/620) | 18.7%<br>(116/620) | 70%<br>(434/620) | -- |  | -- |  | 23.7%<br>(147/620) | 72.3%<br>(448/620) |
| lab_name | 39.7%<br>(140/353) | 36.5%<br>(129/353) | 55.2%<br>(195/353) | 38.8%<br>(137/353) | -- |  | -- |  | 60.1%<br>(212/353) | 38.8%<br>(137/353) |
| specimen_type | -- |  | 90.7%<br>(107/118) | 5.9%<br>(7/118) | -- |  | -- |  | 90.7%<br>(107/118) | 5.9%<br>(7/118) |
| condition | -- |  | 16.8%<br>(16/95) | 66.3%<br>(63/95) | -- |  | 0%<br>(0/95) | 25.3%<br>(24/95) | 16.8%<br>(16/95) | 66.3%<br>(63/95) |
| cause | -- |  | 34.1%<br>(31/91) | 59.3%<br>(54/91) | 3.3%<br>(3/91) | 9.9%<br>(9/91) | 7.7%<br>(7/91) | 34.1%<br>(31/91) | 34.1%<br>(31/91) | 59.3%<br>(54/91) |
| age_group | 14.5%<br>(11/76) | 77.6%<br>(59/76) | 13.2%<br>(10/76) | 84.2%<br>(64/76) | -- |  | -- |  | 14.5%<br>(11/76) | 84.2%<br>(64/76) |
| action | 1.4%<br>(1/71) | 98.6%<br>(70/71) | 0%<br>(0/71) | 80.3%<br>(57/71) | -- |  | -- |  | 1.4%<br>(1/71) | 98.6%<br>(70/71) |
| gender | 5.6%<br>(3/54) | 85.2%<br>(46/54) | 14.8%<br>(8/54) | 85.2%<br>(46/54) | -- |  | -- |  | 14.8%<br>(8/54) | 85.2%<br>(46/54) |

#### Knowledgebase Graph in Neo4j

We stored the annotated data in Neo4j, a graph database management system designed to store, manage, and query graph data. It is a powerful database system that is designed to handle complex, highly connected data. The nodes represent entities, and the relationships represent the connections between those entities. Each node and relationship can have properties, which are key-value pairs that store additional information about the node or relationship. Neo4j also supports graph queries, which allow you to query the graph data using the Cypher query language. Figure 5 shows a graph in Neo4j representing the annotation information for prostate-specific antigen test result.

**Figure 5.**
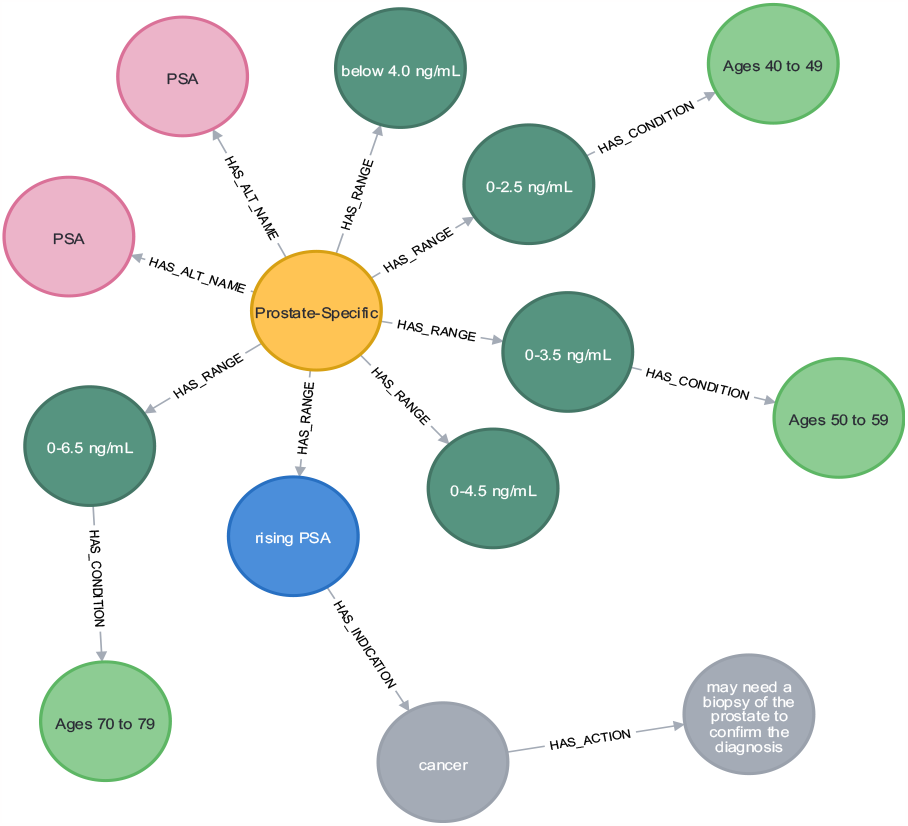
Neo4j subgraph for prostate-specific antigen.

## Discussion and Conclusions

In this study, we collected and annotated result interpretation records from 251 consumer-friendly articles about laboratory tests from AHealthyMe.com. Then we evaluated transformer-based language models for named entity recognition of the key terms and their entity types. We also mapped the annotated terms to concepts in existing controlled terminologies. This work lays the foundation of enhancing patient portals to provide tailored information support for lab result interpretation based on a knowledge graph with information extracted from credible health sources. There are a number of ways in which such a knowledge graph can enhance lab result reporting in patient portals. First, it could provide tailored information support to patients based on their demographics and medical context. For example, prostate-specific antigen (PSA) is a test that measures the protein produced by normal, as well as malignant, cells of the prostate gland. The blood level of PSA is often elevated in people with prostate cancer [25]. As shown in **Figure 5**, the reference range of PSA varies by age and enhanced patient portals can report age-specific reference range and provide possible indications for abnormal results (e.g., prostate cancer) with a follow-up suggested action (i.e., need a biopsy of the prostate).

Patient-centered care highlights the importance of empowering patients to become more proactive in their healthcare and to make more informed decisions. Doctor-patient communication is a key element of patient-centered care and is particularly important for facilitating shared decision-making and for establishing a therapeutic alliance between patients and their health care providers [26]. It influences the quality of patient care and health outcomes, as well as patient motivation, satisfaction, and treatment adherence. Communication skills such as asking/answering questions, listening attentively, sharing critical health information, and providing tailored guidance are all important. The quality and extent of information exchanged during patient encounters, however, can vary based on different factors such as patients’ ages, communication skills, education, health literacy, and personality traits, among others [27]. Previous studies have found that patients with low health literacy have difficulty in formulating questions during physician consult, it is important to prepare patients with a question prompt list.

Based on the previous studies about lab result comprehension [28], annotated comprehension of lab results in this study, and the discussion with MD co-authors, we have identified three types of questions about the reasons for abnormal results: a) procedure-related questions (e.g., “*Is my abnormal result due to a recent surgery?*”), b) medical condition-related questions (e.g., “*Is my abnormal result of creatinine related to my liver disease?*”), c) medication-related questions (e.g., “*Is my abnormal coagulation test result due to use of heparin?*”). As patients with limited health literacy may find it hard to construct a contextualized question about lab results, we can suggest possible questions they can ask during physician consults. Specifically, the questions can be generated based on the user’s EHR data. For example, given the lab test result “albumin: 7.3 g/dL”, we will determine that it is higher than the normal range (3.4 to 5.4 g/dL) based on the Neo4j subgraph of albumin (**Figure 6**). Based on the graph, high albumin level may be caused by “acute infection, burns, and stress from surgery”, we can search for such information in the patient’s EHR. In case the patient had surgery, we can generate questions such as “*Is my low albumin due to stress from surgery?*” – questions that they can discuss with their doctor.

**Figure 6.**
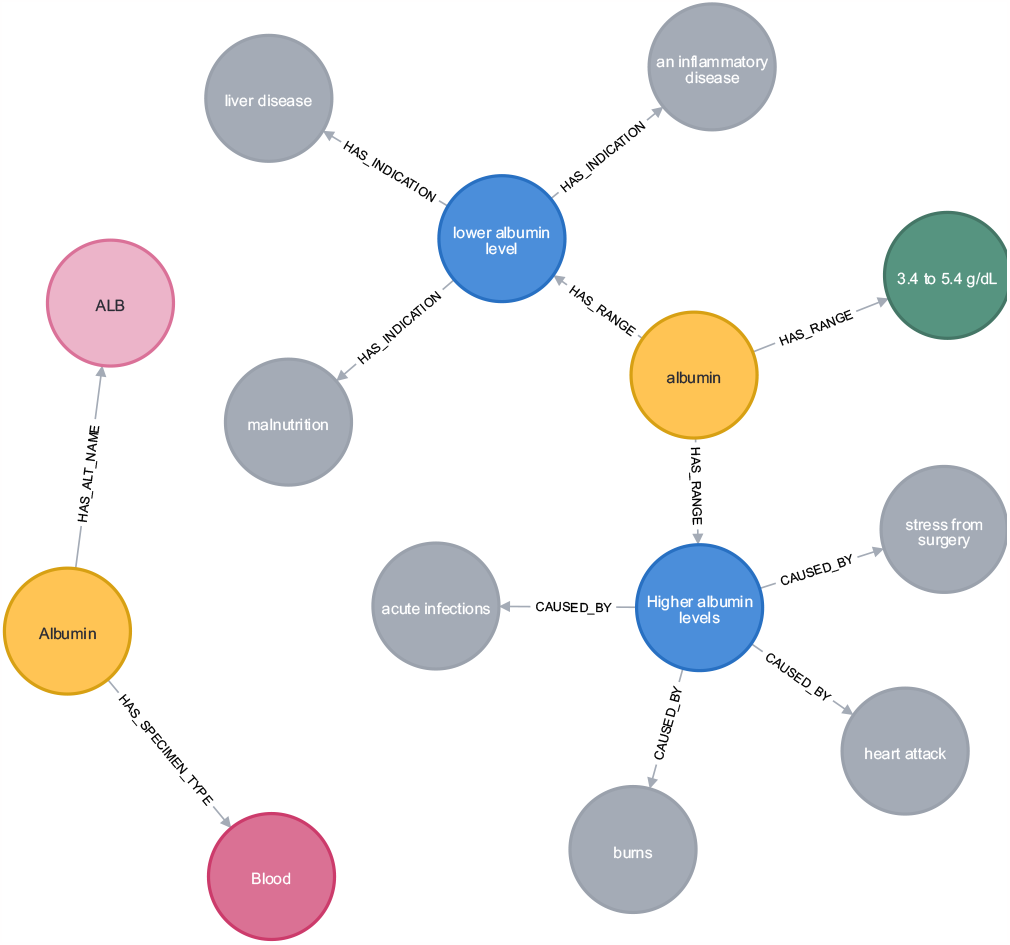
Annotation of lab result interpretation for albumin.

### Limitations

A few limitations need to be noted in this work. First, we used a single corpus collected from AHealthyMe.com for both annotation and evaluation of NER models. Ideally, an external dataset should be used to evaluate the external validity of the models. Second, certain annotations can be refined or broken down into multiple terms (e.g., normal range can be broken down to value range and units).

### Future Work

In future work, we will evaluate models for relationship extraction and develop algorithms and a tool to generate tailored information support and question prompt lists for patients to prepare for their doctor visits. This tool can be linked to patients’ EHR data to provide tailored support and consequently enhance patient understanding and ability to engage in shared decision making with their doctors.

## Data Availability

All data produced in the present study are available upon reasonable request to the authors.

## Acknowledgements

We would like to thank four undergraduate students Jessica Valyou, David Garner, Shawtah Thomas, and Madelyn Dupuis for annotating the data. This work was supported by the Planning Grant of Florida State University Institute for Successful Longevity, National Library of Medicine grant R21LM013911, and in part by the University of Florida – Florida State University Clinical and Translational Science Award UL1TR001427 supported by National Center for Advancing Translational Sciences.

## References

1. Tsai R, Bell EJ, Woo H, Baldwin K, Pfeffer MA. How Patients Use a Patient Portal: An Institutional Case Study of Demographics and Usage Patterns. Appl Clin Inform. 2019 Jan;10(1):96–102.

2. Tapuria A, Porat T, Kalra D, Dsouza G, Xiaohui S, Curcin V. Impact of patient access to their electronic health record: systematic review. Inform Health Soc Care. 2021 Jun 2;46(2):192–204.

3. Kruse CS, Bolton K, Freriks G. The effect of patient portals on quality outcomes and its implications to meaningful use: a systematic review. J Med Internet Res. 2015 Feb 10;17(2):e44.

4. Witteman HO, Zikmund-Fisher BJ. Communicating laboratory results to patients and families. Clin Chem Lab Med. 2019 Feb 25;57(3):359–64.

5. Turchioe MR, Myers A, Isaac S, Baik D, Grossman LV, Ancker JS, et al. A Systematic Review of Patient-Facing Visualizations of Personal Health Data. Appl Clin Inform. 2019 Aug;10(4):751–70.

6. Alpert JM, Krist AH, Aycock RA, Kreps GL. Applying Multiple Methods to Comprehensively Evaluate a Patient Portal’s Effectiveness to Convey Information to Patients. J Med Internet Res. 2016 May 17;18(5):e112.

7. Zikmund-Fisher BJ, Exe NL, Witteman HO. Numeracy and Literacy Independently Predict Patients’ Ability to Identify Out-of-Range Test Results. J Med Internet Res. 2014 Aug 8;16(8):e187.

8. Bar-Lev S, Beimel D. Numbers, graphs and words – do we really understand the lab test results accessible via the patient portals? Isr J Health Policy Res [Internet]. 2020;9(1). Available from: https://www.embase.com/search/results?subaction=viewrecord&id=L2007099289&from=export

9. Pillemer F, Price RA, Paone S, Martich GD, Albert S, Haidari L, et al. Direct Release of Test Results to Patients Increases Patient Engagement and Utilization of Care. PloS One. 2016;11(6):e0154743.

10. Giardina TD, Baldwin J, Nystrom DT, Sittig DF, Singh H. Patient perceptions of receiving test results via online portals: a mixed-methods study. J Am Med Inform Assoc JAMIA. 2018 Apr 1;25(4):440–6.

11. Zhang Z, Kmoth L, Luo X, He Z. User-Centered System Design for Communicating Clinical Laboratory Test Results: Design and Evaluation Study. JMIR Hum Factors. 2021 Nov 25;8(4):e26017.

12. Tao D, Yuan J, Qu X, Wang T, Chen X. Presentation of Personal Health Information for Consumers: An Experimental Comparison of Four Visualization Formats. In: Harris D, editor. Engineering Psychology and Cognitive Ergonomics. Cham: Springer International Publishing; 2018. p. 490–500. (Lecture Notes in Computer Science).

13. Zhang Z, Lu Y, Kou Y, Wu DTY, Huh-Yoo J, He Z. Understanding patient information needs about their clinical laboratory results: A study of social Q&A site. In: 17th World Congress on Medical and Health Informatics, MEDINFO 2019, August 25, 2019 - August 30, 2019. Lyon, France: IOS Press BV; 2019. p. 1403–7. (Studies in Health Technology and Informatics; vol. 264).

14. Kopanitsa G. Study of patients’ attitude to automatic interpretation of laboratory test results and its influence on follow-up rate. BMC Med Inform Decis Mak. 2022 Mar 27;22(1):79.

15. Zikmund-Fisher B, Scherer A, Witteman H, Solomon J, Exe N, Tarini B, et al. Graphics help patients distinguish between urgent and non-urgent deviations in laboratory test results. J Am Med Inform Assoc. 2017 May;24(3):520–8.

16. Zikmund-Fisher BJ, Scherer AM, Witteman HO, Solomon JB, Exe NL, Fagerlin A. Effect of Harm Anchors in Visual Displays of Test Results on Patient Perceptions of Urgency About Near-Normal Values: Experimental Study. J Med Internet Res. 2018 Mar 26;20(3):e98.

17. Scherer AM, Witteman HO, Solomon J, Exe NL, Fagerlin A, Zikmund-Fisher BJ. Improving the Understanding of Test Results by Substituting (Not Adding) Goal Ranges: Web-Based Between-Subjects Experiment. J Med Internet Res. 2018;20(10):e11027.

18. Morrow D, Azevedo RFL, Garcia-Retamero R, Hasegawa-Johnson M, Huang T, Schuh W, et al. Contextualizing numeric clinical test results for gist comprehension: Implications for EHR patient portals. J Exp Psychol Appl. 2019;25(1):41–61.

19. Devlin J, Chang MW, Lee K, Toutanova K. BERT: Pre-training of Deep Bidirectional Transformers for Language Understanding [Internet]. arXiv; 2019 [cited 2023 Mar 6]. Available from: http://arxiv.org/abs/1810.04805

20. Lee J, Yoon W, Kim S, Kim D, Kim S, So CH, et al. BioBERT: a pre-trained biomedical language representation model for biomedical text mining. Bioinformatics. 2020 Feb 15;36(4):1234–40.

21. Huang K, Altosaar J, Ranganath R. ClinicalBERT: Modeling Clinical Notes and Predicting Hospital Readmission [Internet]. arXiv; 2020 [cited 2023 Mar 6]. Available from: http://arxiv.org/abs/1904.05342

22. Liu Y, Ott M, Goyal N, Du J, Joshi M, Chen D, et al. RoBERTa: A Robustly Optimized BERT Pretraining Approach [Internet]. arXiv; 2019 [cited 2023 Mar 6]. Available from: http://arxiv.org/abs/1907.11692

23. Tian S, Erdengasileng A, Yang X, Guo Y, Wu Y, Zhang J, et al. Transformer-Based Named Entity Recognition for Parsing Clinical Trial Eligibility Criteria. ACM-BCB ACM Conf Bioinforma Comput Biol Biomed ACM Conf Bioinforma Comput Biol Biomed. 2021 Aug;2021:49.

24. Gu Y, Tinn R, Cheng H, Lucas M, Usuyama N, Liu X, et al. Domain-Specific Language Model Pretraining for Biomedical Natural Language Processing. ACM Trans Comput Healthc. 2021 Oct 15;3(1):2:1-2:23.

25. Prostate-Specific Antigen (PSA) Test - NCI [Internet]. 2022 [cited 2023 Mar 7]. Available from: https://www.cancer.gov/types/prostate/psa-fact-sheet

26. Rathert C, Wyrwich MD, Boren SA. Patient-centered care and outcomes: a systematic review of the literature. Med Care Res Rev MCRR. 2013 Aug;70(4):351–79.

27. Keinki C, Momberg A, Clauß K, Bozkurt G, Hertel E, Freuding M, et al. Effect of question prompt lists for cancer patients on communication and mental health outcomes-A systematic review. Patient Educ Couns. 2021 Jun;104(6):1335–46.

28. Zhang Z, Citardi D, Xing A, Luo X, Lu Y, He Z. Patient Challenges and Needs in Comprehending Laboratory Test Results: Mixed Methods Study. J Med Internet Res. 2020 Dec 7;22(12):e18725.

